# Validation of the Baseline Recurrence Risk in Cellulitis (BRRISC) score and the added impact of acute clinical response

**DOI:** 10.1101/2024.12.18.24319216

**Authors:** Elizabeth LA Cross, Gail N Hayward, Martin J Llewelyn, A Sarah Walker

## Abstract

**Objectives:** The BRRISC score predicts hospital-attended recurrence of cellulitis using baseline clinical data. In practice, clinicians assess treatment response when deciding antibiotic duration. We investigated whether markers of acute clinical response improved prognostic score performance within an external validation cohort.

**Methods:** We recruited adults with lower limb cellulitis attending hospital. Outcomes included ‘hospital-attended recurrence’ (primary outcome for validation) and ‘any recurrence’. We assessed clinical response from days 0-3 of treatment. Using multivariate logistic regression with backwards elimination, we identified response variables independently associated with either outcome for an extended score.

**Results:** Of 202 patients, 8% (n=17) experienced ‘hospital-attended recurrence’ and 23% (n=46) ‘any recurrence’. In this validation dataset, the BRRISC score had a C-index=0.75 (95%CI, 0.64-0.86) for predicting ‘hospital-attended recurrence’. There was weak evidence that an extended score, incorporating day-2/3 severity of skin blistering, improved the C-index to 0.83 (0.75-0.92). No acute clinical response variables were independently associated with ‘any recurrence’ after adjusting for BRRISC score.

**Conclusion:** Our measures of early clinical response did not add useful prognostic information to BRRISC score baseline risk. Further research should consider whether responses after day-3 add helpful information, whether risk stratification can personalise antibiotic duration, and explore non-antibiotic approaches to prevent recurrence.

## Introduction

Cellulitis is a common and debilitating skin infection characterised by pain, swelling, tenderness, increased temperature, and acute colour change of the affected area.^1^ Although most patients recover clinically in the short-term, 15-50% experience recurrence in the months following an initial episode, resulting in repeated consultations and treatment.^2-5^ Consequently, cellulitis recurrence is the outcome most frequently identified as important to patients and healthcare professionals.^6^

Treatment guidelines for cellulitis recommend 5-7 days of antibiotics,^7,8^ but evidence demonstrates that most patients are treated for considerably longer.^9-11^ Clinicians may prolong therapy to try to avoid poor clinical outcomes but, in doing so, frequently overtreat patients. Unnecessary antibiotic use harms patients through antibiotic adverse effects and resistant infections.^12^ Whether prolonged antibiotic treatment reduces recurrence risk in some patients is unclear.^4^

We previously developed the BRRISC score, which stratifies patients as low (score 0-1), medium (2-5), or high (6-15) risk of experiencing hospital-attended cellulitis recurrence.^13^ The score was derived using electronic health records (EHRs) containing clinical data available at baseline (age, heart rate, urea, platelet count, albumin) and comorbidities (previous cellulitis, venous insufficiency, liver disease). While recurrence rates increased fourfold across risk groups, score performance was only moderate (C-index=0.65 (95% CI, 0.63-0.68). We hypothesised that this might reflect a lack of data on clinical response, which clinicians use in everyday practice when deciding antibiotic therapy duration.^14,15^

Here, we followed a cohort of UK adults with lower limb cellulitis to capture early clinical response and recurrences. Our objectives were to externally validate the BRRISC score for hospital-attended recurrence, test the score’s performance for predicting any recurrence (hospital-attended or community), and determine whether adding acute clinical response would improve prognostic value.

## Methods

### Population and setting

Patients were recruited from two acute hospitals in the United Kingdom’s National Health Service (NHS): a large tertiary referral hospital and a district general hospital, both within University Hospitals Sussex NHS Foundation Trust, from June 2021 to March 2023. Adults (_≥_18 years) were eligible if their treating clinician identified them as having lower limb cellulitis requiring intravenous or oral antibiotic treatment. The main exclusion criteria were already receiving _≥_3 calendar days of antibiotics from the hospital for cellulitis, being treated for a previous cellulitis episode in the preceding 28 days, or if the clinical diagnosis changed to an alternative diagnosis within three days of original enrolment (**Supplementary Figure 1;** details in **Appendix p1**). The population on which the BRRISC score was originally developed differed in that patients with cellulitis at sites other than the lower limb (but excluding facial cellulitis) were also included due to difficulties with accurately defining lower limb cellulitis from diagnostic codes in EHRs.^13^ See **Appendix p2-3** for sample size calculation.

### Study assessments

Data was collected on demographics, comorbidities (including details of current and previous cellulitis episodes), microbiological results, and antibiotic use. ‘Previous cellulitis’ was self-reported by patients. To replicate the definition of ‘previous cellulitis’ used in BRRISC score development, ‘hospital-attended previous cellulitis’ was defined as any previous hospital attendance containing a cellulitis diagnosis code in any position identified from EHRs (**Appendix p3**).

Acute clinical responses were assessed by physical examination (including the use of a Cellulitis Severity Score (CSS)^4,16^, **Supplementary Table 1**), limb temperature measurements, vital signs, blood test results (infection markers (C-reactive protein (CRP), neutrophil count) and tests from the BRRISC score (platelets, urea, albumin)), and patient-reported pain and swelling due to cellulitis. Where possible, temperature measurements were performed on all patients daily for four days beginning on day-0, defined as the date the patient began their hospital-associated antibiotic treatment for cellulitis. Where patients were enrolled after day-0, physical examination and temperature readings were only available from enrolment. Where patients were discharged before day-3, readings were only available until discharge. We did not perform additional blood tests for the study but used those available from clinical care. For further details on acute clinical response variables, see **Appendix p3**. We used mixed models to estimate changes in these parameters over time, assuming data were missing at random (correlated patient-specific intercept and slope).

### Outcomes

The primary outcome was ‘any recurrence’ within 90 days, defined as initiating new antibiotic treatment in primary care or hospital for cellulitis at the same site. Recurrence was assessed at a follow-up visit or if the patient was uncontactable via EHRs. The BRRISC score was originally developed using the narrower definition of ‘hospital-attended recurrence’ within 90 days, so this was also considered for primary score validation.

### Statistical analysis

We truncated vital signs and blood results at the 1st and 99th percentiles to avoid undue influence from outliers. For missing data in acute clinical responses, we considered both hard imputations (day 0/1=day-0 otherwise day-1; day 2/3=day-2 otherwise day-3) and performed multiple imputation of variables in **Supplementary Table 2** by chained equations with predictive mean matching (missing at random assumption; 50 imputed datasets). The 40 imputed variables were those in the BRRISC score and acute clinical response variables (absolute and change from baseline) with univariate p<0.15 for either recurrence outcome. Imputed and observed values were compared visually (**Supplementary Figures 2, 3**).

We first externally validated the BRRISC score’s ability to predict ‘hospital-attended recurrence’, the outcome for which it was developed. Given patients’ age and co-morbidities, and the fact that risk factors for death are likely to be very different to those for recurrence, those who died within 90 days before experiencing a recurrence were excluded from the analysis (effectively considering the first two components of a multinomial outcome; no recurrence/death, recurrence before death, death before recurrence), as was done for original score development.^13^ We subsequently tested the score’s ability to predict the new outcome, ‘any recurrence’. Performance was measured using Harrell’s C-index/area under the ROC curve (AUC) in the imputed datasets, with values <0.6 considered poor, 0.6-0.7 moderate, and >0.7 good.^17,18^ For each BRRISC score level, mean observed risks across imputed datasets were compared versus predicted risks from the original model, combining score levels where some imputations contained zero recurrences or only recurrences in one or more of the score levels. Similarly to the outcomes, the BRRISC score variable ‘previous cellulitis’ can be defined as ‘patient-reported’ or ‘hospital-attended’, so the above steps were performed for each definition, leading to the evaluation of four separate prediction models.

### Score extension

To identify whether acute clinical response could improve BRRISC score performance, we considered all additional variables with univariate p<0.05 for either recurrence outcome (details in **Supplementary Table 3**) alongside the BRRISC score in multivariate logistic regression models, using backwards elimination on non-BRRISC score variables to identify independent predictors, and using fractional polynomials to allow for non-linearity in continuous factors. We used a relatively strict univariate threshold to favour a parsimonious model, given this was an extension of the baseline risk model. Beta-coefficients of independent predictors were used to calculate the number of additional score points that should be added to the BRRISC score. We compared the C-indexes from the original and extended scores in each imputed dataset,^19^ and assessed the contribution of new predictors using a category-based Net Reclassification Index (NRI), which evaluates the proportion of subjects moving accurately or inaccurately from one risk category to another. Three risk categories based on predefined 5% and 15% thresholds were used,^13^ calculating event and non-event NRIs across the full dataset of multiple imputations. Large positive values indicate that the extended score correctly increases risk estimates for those with recurrences or decreases risk estimates for those without recurrences for event and non-event NRIs, respectively.

Stata v18.0 software (SPSS Inc., Chicago, Illinois) was used for all analyses. We followed the Transparent Reporting of a multivariable prediction model for Individual Prognosis Or Diagnosis (TRIPOD) statement (**Supplementary Table 4**).^20^

### Public and Patient Involvement

This study involved patients and the public in the design and conduct of the research through the James Lind Alliance Cellulitis Priority Setting Partnership^21^ and a patient and public involvement (PPI) group consisting of people with lived experience of cellulitis. Our PPI contributors checked the acceptability of the study procedures, edited patient information materials, and improved outcome measure collection.

### Ethics

This study was approved by the East of Scotland Research Ethics Service, Research Ethics Committee (21/ES/0048). All participants provided written informed consent.

## Results

202 patients with lower limb cellulitis were included in the 90-day recurrence analysis (**Supplementary Figure 1**); the median age was 66 years (interquartile range, IQR 51,79), 84 (42%) were female, and 191 (95%) of white ethnicity. Over half (105, 52%) self-reported a previous cellulitis episode, with 55 (27%) a previous episode requiring hospital attendance, higher than that of the development cohort (5%) (further patient characteristics in **Supplementary Table 5**).

### Acute clinical response

Over days 0-3, total CSS, affected limb temperature, respiratory rate, heart rate, body temperature, CRP, neutrophil count, urea, albumin, and patient-reported symptoms decreased (P<0.001), whilst platelet count increased (P<0.001) (observed data and results from linear mixed models **Figure 1**). There was moderate evidence for decreases in limb temperature difference (P=0.04), weak evidence for decreases in affected skin area (P=0.051), and no evidence for changes in systolic blood pressure (P=0.27).

**Figure 1.**
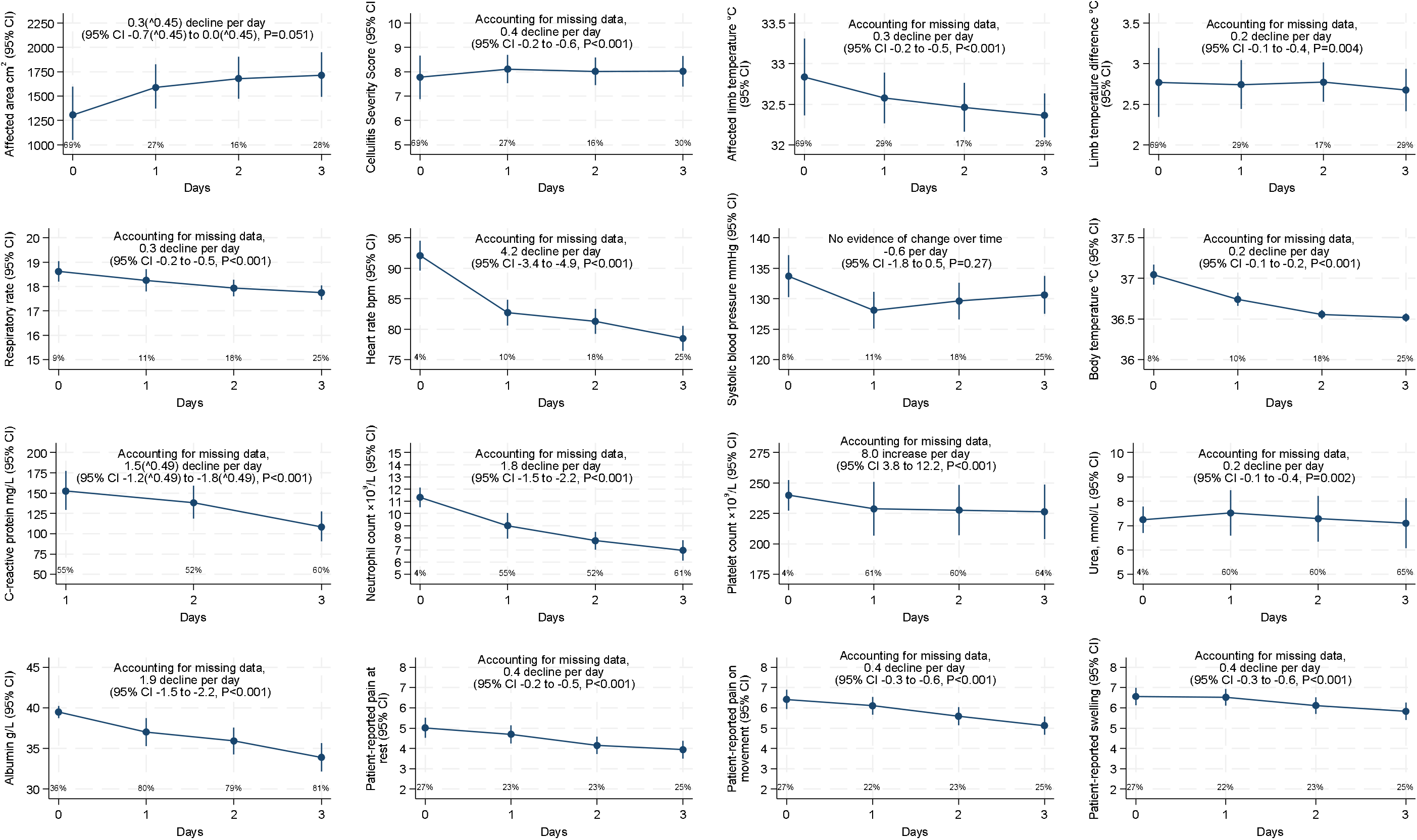
Acute clinical responses over first four days from initiation of antibiotics (Day-0) Note: Day-0 was taken as the date of hospital antibiotic initiation. Percentages indicate missing data per day. Points show observed data. Estimates of change over time from linear mixed models with correlated patient-specific intercept and slope which allow for missing data at random (which includes missing data depending on previous observed values). Affected area and CRP stated declines are transformed values. CSS (Cellulitis Severity Score): Made up of 7 components, each scored 0-3, higher numbers indicate more severe findings, see **Supplementary Table 1**. Patient-reported symptoms scored 0-10, higher numbers indicate more severe symptoms.

In terms of CSS subcomponents, the odds of higher colour, warmth, and oedema subscore decreased per day by 53% (95%CI 0.34-0.64, P<0.001), 53% (0.37-0.60, P<0.001), and 27% (0.55-0.98, P=0.04), respectively (**Figure 2**). The odds of a higher blistering subscore increased per day by 96% (1.07-3.63, P=0.03). There was no evidence for a change in the tenderness subscore over time (OR=0.86, 0.67-1.10, P=0.24) (models for ulceration and discharge subscores did not converge).

**Figure 2.**
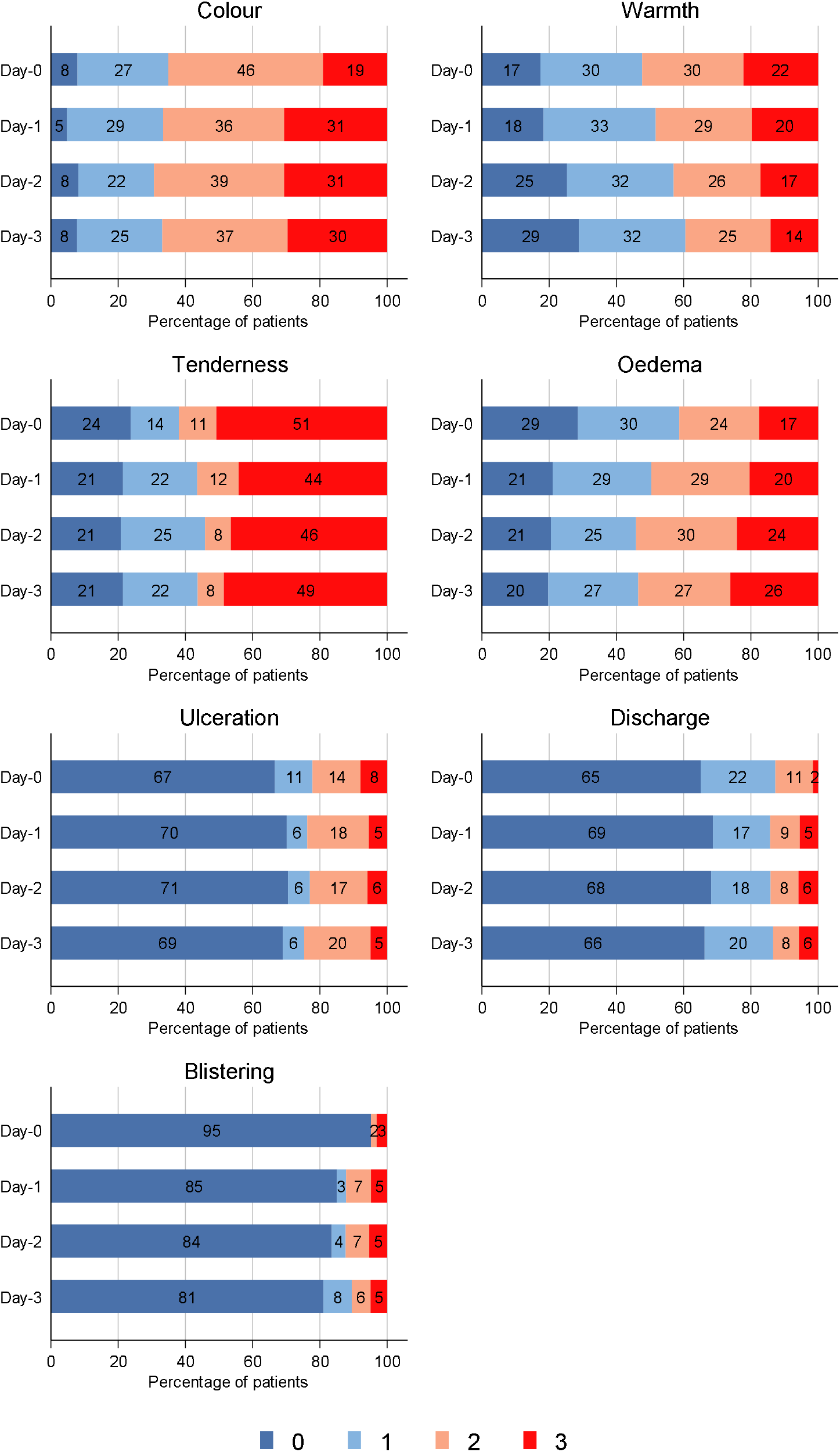
Cellulitis Severity Score (CSS) subcomponents over first four days from initiation of antibiotics (Day-0) Note: Day-0 was taken as the date of hospital antibiotic initiation. The missing data for days 0-3 were 69%, 27%, 16%, and 30%, respectively.

### Antibiotic use

Before hospital attendance, 61 (30%) patients received a median 3 days (IQR 2,4) oral antibiotic therapy from the community. The most common hospital-prescribed antibiotics were flucloxacillin (85,42%), ceftriaxone (52,26%), and clindamycin (28,14%). The initial antibiotic prescribed was most frequently ceftriaxone (91,45%), followed by flucloxacillin (75,37%), and co-amoxiclav (13,6%). All other agents were prescribed to <5% of patients. The median antibiotic duration/length of therapy was 13 days (IQR 10,18), and antibiotic days of therapy 15 days (IQR 10,22) (details in **Supplementary Table 5**).

### Outcomes

The primary study outcome, ‘any recurrence’ within 90 days, occurred in 46 (23%) patients, a median of 31 (IQR 22,56) days from initial hospital attendance (**Supplementary Figure 4**). ‘Hospital-attended recurrence’ (primary outcome for BRRISC validation) occurred in 17 (8%) patients, a median 37 (IQR 24,66) days from initial hospital attendance.

### BRRISC score external validation and test

The BRRISC score had a C-index of 0.75 (95%CI, 0.64-0.86) for predicting ‘hospital-attended recurrence’, regardless of the definition of previous cellulitis used (**Table 1**) vs 0.65 (0.63-0.68) in the original development population based on EHRs.^13^ When tested on the new outcome ‘any recurrence’, the C-index was 0.60 (0.51-0.69) and 0.63 (0.54-0.71) for ‘patient-reported’ or ‘hospital-attended’ definitions of previous cellulitis, respectively.

**Table 1.** Model performance.

| Definition of previous cellulitis | Variable | Beta Coefficient (per unit higher) | OR | 95%CI | P | AUC | 95%CI |
| --- | --- | --- | --- | --- | --- | --- | --- |
| External validation of BRRISC score (original score and outcome (hospital-attended recurrence)) |  |  |  |  |  |  |  |
| Patient-reported | BRRISC score | 0.43 | 1.54 | 1.19-2.00 | 0.001 | 0.75 | 0.65-0.85 |
| Hospital-attended | BRRISC score | 0.42 | 1.52 | 1.19-1.94 | 0.001 | 0.75 | 0.64-0.86 |
| Test of BRRISC score with new outcome (any recurrence) |  |  |  |  |  |  |  |
| Patient-reported | BRRISC score | 0.17 | 1.19 | 1.00-1.41 | 0.04 | 0.60 | 0.51-0.69 |
| Hospital-attended | BRRISC score | 0.21 | 1.24 | 1.04-1.46 | 0.01 | 0.63 | 0.54-0.71 |
| Model including BRRISC score and day-2/3 blistering (original outcome (hospital-attended recurrence)) |  |  |  |  |  |  |  |
| Patient-reported | BRRISC score | 0.52 | 1.68 | 1.25-2.26 | 0.001 | 0.84 | 0.75-0.92 |
|  | Day-2/3 blistering* | 0.97 | 2.63 | 1.56-4.43 | <0.001 |  |  |
| Hospital-attended | BRRISC score | 0.50 | 1.65 | 1.24-2.20 | 0.001 | 0.82 | 0.72-0.93 |
|  | Day-2/3 blistering* | 0.95 | 2.58 | 1.55-4.29 | <0.001 |  |  |
| Extended score (incorporating day-2/3 blistering into score (original outcome (hospital-attended recurrence)) |  |  |  |  |  |  |  |
| Patient-reported | Extended score | 0.40 | 1.49 | 1.24-1.79 | <0.001 | 0.83 | 0.75-0.92 |
| Hospital-attended | Extended score | 0.39 | 1.48 | 1.24-1.76 | <0.001 | 0.82 | 0.72-0.92 |

\*Blistering included as linear; see **Supplementary Table 6** for model including blistering as categorical.
Note: In development dataset, using hospital-attended previous cellulitis and hospital-attended recurrence outcome, AUC=0.65 (95% CI, 0.63-0.68) (validation equivalent shown in bold).<sup>13</sup>

The percentages with recurrence by either previous cellulitis definition generally increased with increasing BRRISC score up to a score of 6, with wide 95% CI above this (**Figure 3**). For the ‘hospital-attended recurrence’ outcome, on which the score was developed, 95% CIs generally overlapped the risk predictions from the original model, whereas risk estimates were generally higher for the ‘any recurrence’ outcome, as expected.

**Figure 3.**
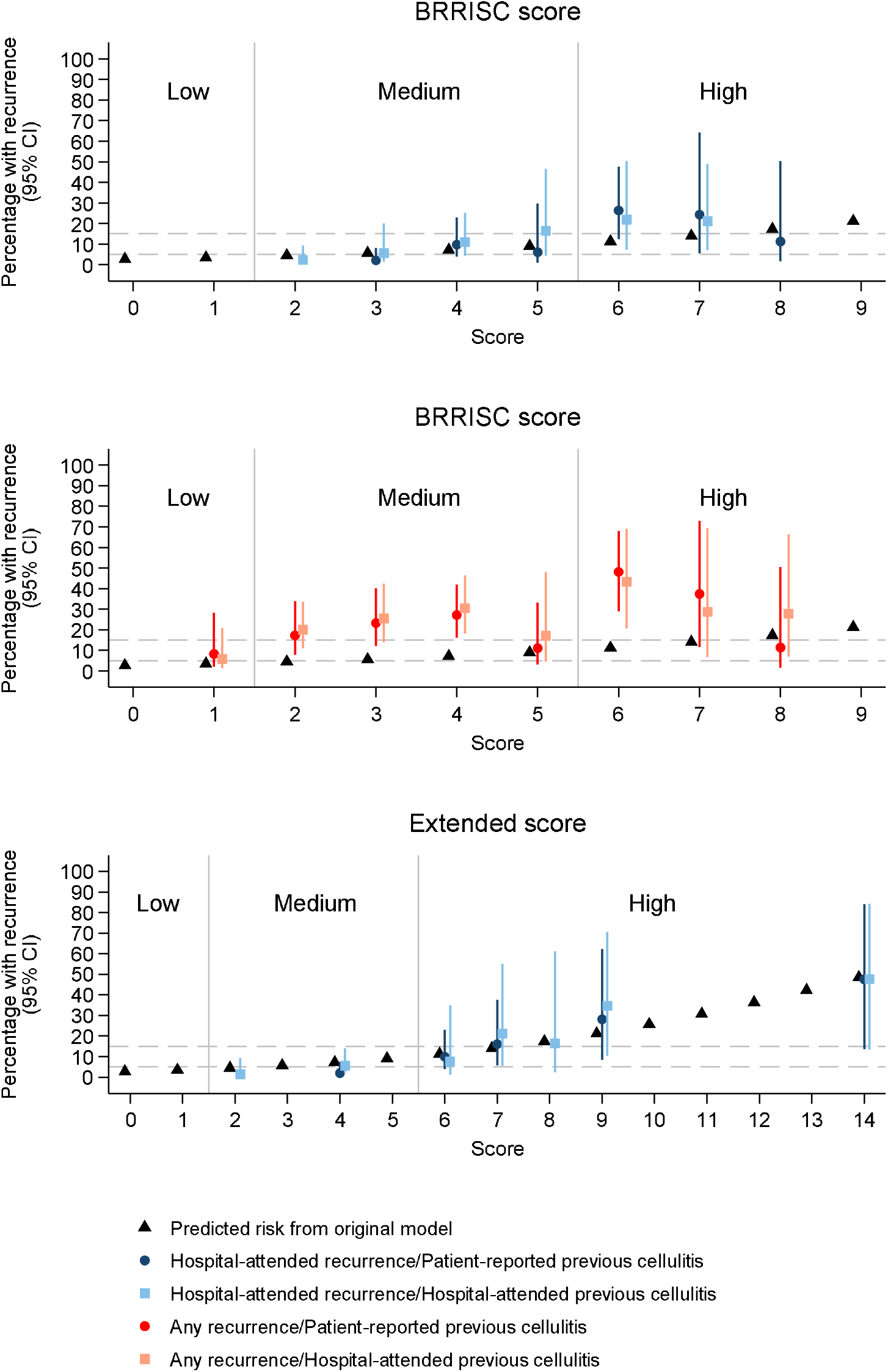
Observed (95%CI) and predicted percentage with recurrence per score level. Note: As some imputations contained zero recurrences in one or more of the score levels, observed risks were combined across some score levels to calculate estimated observed risk across imputed datasets (BRRISC score dark blue 3=0-3, 8=8-9, light blue 2=0-2, 7=7-9, red and pink 1=0-1, 8=8-9; Extended score dark blue 4=0-4, 6=5-6 7=7-8, 9=9-11, 14=12-14 & 17, light blue 2=0-2, 4=3-5, 9=9-11, 14=12-14 & 17). Previously defined risk thresholds at 5% and 15% categorised scores as low (score 0-1), medium (2-5) and high (6-15) risk. Supplementary Table 7 for Net Reclassification Index relating to extended score.

### BRRISC score extension using acute clinical response

Of the acute clinical response variables univariately associated with ‘hospital-attended recurrence’ (**Supplementary Table 3**), only day-2/3 skin blistering (part of CSS, see **Supplementary Table 1**) was independently associated after adjusting for the BRRISC score, with C-indexes 0.84 (95%CI, 0.75-0.92) and 0.82 (0.72-0.93) for ‘patient-reported’ and ‘hospital-attended’ previous cellulitis definitions, respectively (**Table 1**). Assigning 2, 4, and 8 points for day-2/3 blistering subscores of 1, 2, and 3, respectively, based on beta coefficients from the combined model (considering day-2/3 blistering linearly (**Table 1**) or categorically (**Supplementary Table 6**)) increased the maximum possible score from 15 to 23 points, whilst maintaining extended model performance (**Table 1, Figure 3)**. Tests of equality of the BRRISC and extended score C-indexes in each imputed dataset showed most differences between C-indexes were >0.075 (N=31), and most P-values <0.15 (N=32), indicating weak evidence that the extended score C-index was higher than the BRRISC score (**Supplementary Figures 5&6**).

Using the extended BRRISC score, the reclassification of imputations into risk groups produced an event NRI of +33%/+16% (correctly reclassified high-risk patients) and non-event NRI of -12%/-3% (more often incorrectly classified low-risk patients as high-risk), using ‘patient-reported’/’hospital-attended’ previous cellulitis, respectively (**Table 3**). The summary NRIs were, therefore, +22% and +13%, respectively.

As none of the variables univariately associated with the new ‘any recurrence’ outcome at p<0.15 (**Supplementary Table 3**) remained associated with the outcome at p<0.05 after adjusting for the BRRISC score, we did not consider extending the BRRISC score to predict ‘any recurrence’.

## Discussion

We have confirmed the performance of the BRRISC score in predicting hospital-attended recurrence of cellulitis in a prospective cohort of UK adults. Although patients experienced improvements in the acute clinical response to infection over the first four days of treatment, we found weak evidence that including markers of treatment response into the BRRISC score improved its performance. Moreover, the only marker that improved the score’s performance was the level of blistering on day-2/3, which was arguably more likely to reflect the severity of the initial illness rather than a positive response to treatment. Of note, none of the acute clinical response variables independently predicted ‘any recurrence’ after adjusting for the BRRISC score. It is possible that this is because patients with severe blistering may experience equally severe recurrences requiring hospital admission.

Extending the score to include day-2/3 blistering more often incorrectly classified low-risk patients as high-risk, possibly because the additional points did not always accurately reflect risk level (i.e. 8 points from day-2/3 blistering automatically categorised a patient as high-risk, even if they possessed no other risk factors). Therefore, and because the evidence for its added prognostic value is relatively weak, we would not recommend adding day-2/3 blistering to the BRRISC score.

A potential limitation of our study is that by only assessing response up to day-3, we could have missed the potential prognostic value of assessment after that time point. Willliams *et al*. examined data from the adjunctive clindamycin for cellulitis clinical trial and described a delay in the resolution of local symptoms and signs.^22^ For instance, by day-10, 36.4% of individuals still had a _≥_2 cm difference in circumference, and 29.5% had _≥_1°C difference in skin temperature between affected and unaffected limbs. However, they did find very strong evidence for a decrease in these variables by day-5 (median 4 days follow-up). The only other study to monitor limb temperature over time reported significant decreases in temperature change between the affected and unaffected limbs during the first 3 days of treatment, but no evidence for decreases from day-4 until day-7, although the sample size was smaller during this period.^23^ Current guidelines recommend basing antibiotic decision-making on a review of treatment response at 48-72 hours.^8,24,25^

In a recent pilot study testing an algorithm for intravenous to oral switch in 128 hospitalised patients with cellulitis, just over three-quarters showed significant clinical improvement within 48 hours of treatment.^11^ Similarly, among 216 patients with cellulitis, Bruun *et al*. found 90% had a measurable improvement in local inflammation and CRP by day-3 of treatment, and the strongest concordance between clinical and biochemical response occurred on days-2/3.^14^ As in our study, day-3 response did not predict a poor outcome (clinical failure).

A systematic review and meta-analysis of cellulitis of antibiotic treatment trials with data on time to clinical response found the time to clinical response overall was 1.68 days (95% CI, 1.48-1.88).^26^ However, time courses varied between parameters. For example, the proportion of patients with oedema fell 30-50% by day 2-4 and there was a ∼50% reduction in pain and severity scores by day-5. The authors concluded that the optimal clinical reassessment time is likely between 2 to 4 days, but this was based on four heterogeneous studies.

A further limitation is that we used a relatively crude method for measuring change in the size of affected skin area. This could explain why we found only weak evidence for a decrease despite a reduction in lesion size at 48-72 hours being the recommended primary efficacy endpoint for cellulitis antibiotic trials.^27^ Similarly, we measured skin temperature by locating the hottest point on the limb, but more advanced techniques that capture changes in the area of increased temperature are emerging.^28^ Nonetheless, the approach we used was selected because it exemplifies the sort of approach clinicians could realistically apply in everyday practice.

Nearly one-third of our patients had had antibiotics before admission. This is in keeping with other observational studies in similar settings, but assuming some of these patients were presenting with partially treated infection rather than treatment failure,^10^ this could have diluted our ability to detect a link between response to treatment after admission and outcome.

Our pragmatic inclusion criteria of ‘clinician-treated cellulitis’ is another source of potential dilutional bias. If some patients did not have true cellulitis,^29^ they would not be expected to respond to antibiotic treatment. However, we took particular care to exclude patients from our study if the clinical diagnosis changed to an alternative diagnosis within three days of enrolment or if the investigator judged they did not have a clear diagnosis of cellulitis.

Other limitations of the study relate to generalisability. We only included patients from two hospitals, although they represented a mixture of tertiary care and district general services. Most patients were of white ethnicity (the case for most studies in this area), so measurement of clinical response in people with different skin tones, particularly changes in the affected area and skin temperature, needs further exploration. Our findings do not necessarily apply to cellulitis affecting other body parts, less severe cases managed in the community, and/or patients treated with only oral therapy (only 6.4% of our cohort).

Our study also has many strengths. We successfully externally validated the BRRISC score in a new patient population, demonstrating that baseline clinical information related to patient predisposition and illness severity can predict hospital-attended recurrence. This is particularly relevant as hospital-attended recurrences represent more severe disease and are where most treatment costs are incurred.^30^ We found high levels of self-reported and hospital-attended previous cellulitis in our cohort (52% and 27%, respectively), demonstrating that while cellulitis is often viewed as an acute illness, many patients suffer from a chronic burden of recurrent disease.

We assessed a wide range of acute clinical response variables, including physical examination findings, biochemical changes, and patient-reported symptoms. We also used an objective method for monitoring limb temperature over time, which only three studies have previously attempted.^22,23,28^ In terms of outcomes, we observed patients until day-90, while most previous studies’ have followed patients up to _≤_30 days,^31^ allowing us to capture disease recurrences fully. Finally, our findings add to the growing body of evidence that most patients with cellulitis are treated for considerably longer than guidelines recommend.^9-11^

Future studies should explore whether treatment response between day-4 to 7 is prognostic, ensuring measurement approaches are accessible and interpretable in clinical settings in real-time. Controlled trials are required to establish whether risk of recurrence can be used to inform the selection of antibiotic treatment options and, if treatment response after day-3 were predictive, these would need to consider durations longer than the current 5-7 days for at least some patients. Consideration should also be given to non-antibiotic approaches (e.g. management of limb oedema) in patients at high risk of recurrence.^5^

## Data Availability

The data underlying this article will be shared on reasonable request to the corresponding author.

## Data sharing

The data underlying this article will be shared on reasonable request to the corresponding author.

