## Supplementary material for "Validation of the Baseline Recurrence Risk in Cellulitis (BRRISC) score and the added impact of acute clinical response"

### Supplementary materials

[Supplementary Table 2. Univariable associations between acute clinical response variables and ‘hospital-attended recurrence’ or ‘any recurrence’ used to determine which variables to impute (threshold P<0.15)	9](#_Toc185414436)

[Supplementary Table 3. Univariable associations between acute clinical response variables and either ‘hospital-attended’ or ‘any recurrence’ with p<0.05	9](#_Toc185414437)

### Supplementary methods

#### Inclusion and exclusion criteria

Further exclusion criteria were patients who:

- received antibiotic therapy for another indication that was anticipated to continue for longer than the antibiotic treatment for cellulitis and that, in the judgement of the investigator, would have impacted the study assessments.
- required a surgical procedure to treat their infection (i.e. debridement of suspected necrotising skin / soft tissue infection).
- in the judgement of the investigator, did not have a clear diagnosis of cellulitis (to enable the exclusion of infections, such as severe/deep diabetic foot infection, which may be loosely labelled as cellulitis, but treated with different guideline antibiotic agents and durations)
- lacked capacity to give informed consent to participate.
- were receiving end-of-life care.
- were already involved in a clinical trial of an investigational medicinal product (CTIMP) of relevance to the treatment of their cellulitis.
- were unlikely, in the opinion of the investigator, to comply with study procedures.

**Sample size**

As we did not know the prevalence of the different predictors when designing the study, we considered predictors present in the final sample size at prevalence 10%, 20%, 30%... 90%. We assumed that the overall recurrence rate (hospital or community) would be 14%,^1^ and that differences in this event rate between those with and without a predictor of interest would be 15%, and used these to determine the event rates in those with and without a hypothesised predictor giving 14% outcomes overall. We then calculated a standard two-group binomial sample size assuming 80% power and 2-sided alpha=0.05, repeating this for a 20% difference. Assuming 10% loss to follow-up, 220 patients provided 80% power to detect differences in recurrence rates of 15% associated with high-risk exposures prevalent at >30% and of 20% associated with high-risk exposures prevalent at 15-30%.

```
* difference in outcome rate of 15%

noi di _n _dup(80) "*" _n "15% difference in outcome" _n _dup(80) "*"
forval i=0.1(0.1)0.9 {
* proportion in second group
local j=1-`i'
* overall event rate is 0.14: find power to detect 0.15 difference between groups
* so if x is rate in higher risk group with prevalence `i'
* y is rate in lower  risk group with prevalence `j'
* x-y=0.15, then
 local y=0.14-0.15*`i'
 local x=0.15+`y'
 local nratio =`j'/`i'
 noi power twoprop `x' `y', n(200) nratio(`nratio')
}

* difference in outcome rate of 20%

noi di _n _dup(80) "*" _n "20% difference in outcome" _n _dup(80) "*"
forval i=0.1(0.05)0.3 {
   * proportion in second group
   local j=1-`i'
   * overall event rate is 0.14: find power to detect 0.20 difference between     groups
   * so if x is rate in higher risk group with prevalence `i'
   * y is rate in lower  risk group with prevalence `j'
   * x-y=0.20, then
   local y=0.14-0.20*`i'
   local x=0.20+`y'
   local nratio =`j'/`i'
   * can't make this work for all values
   if `y'>0 noi power twoprop `x' `y', n(200) nratio(`nratio')
}
```

#### Comorbidities

We computed Charlson and Elixhauser scores using the “CHARLSON” Stata module,^2^ which uses currently recommended Quan (Charlson)^3^ and van Walraven (Elixhauser)^4,5^ ICD-10 codes and weightings for 17 (Charlson) and 31 (Elixhauser) comorbidities. We incorporated primary and secondary diagnostic codes from a one-year lookback (prior to the index hospital admission date), and secondary diagnostic codes recorded during the index cellulitis hospital episode.^6^ Via the same one-year lookback, we used the following ICD-10 codes to identify specific chronic comorbidities identified as potential predictors i) chronic venous insufficiency: I831, I832, I839, O220, I872; ii) other ulcer disease: L890, L891, L892, L893, L899, L97, L984; iii) lymphoedema: I890, I972, Q820; iv) peripheral neuropathy: G60-G63.8; v) immunosuppression:6 B20–24, O987 (AIDS/HIV), C77-96 (metastatic cancer, haematological malignancies), D80-84 (primary immunodeficiencies), K721, K729, K766, K767 (end-stage liver disease). The comorbidity ‘previous cellulitis’ was determined by patients with a previous hospital attendance containing a cellulitis diagnosis code in any position.

*Cellulitis diagnosis codes:*

A46 Erysipelas

L03.0 cellulitis of finger and toe

L03.1 cellulitis of other parts of limb

L03.3 cellulitis of trunk

L03.8 cellulitis of other sites

L03.9 cellulitis unspecified

L08.8 other specified local infections of skin and subcutaneous tissue

L08.9 local infection of skin and subcutaneous tissue, unspecified

#### Acute clinical response variables

*Physical examination:*

- ‘Area’ of acute colour change, oedema, or induration (whichever was largest) was measured with a flexible disposable tape measure, by multiplying the longest head-to-toe length of the lesion with the widest width perpendicular to that length
- A Cellulitis Severity Score (CSS) previously proposed as a descriptive symptom score (not validated, **Supplementary Table 1**).^1,7^ To minimise inter-observer variation for the score, the Chief Investigator performed this assessment in addition to another member of the research team until assessments performed independently matched consistently.

*Vital signs:*

- Vital signs included respiratory rate (RR), heart rate (HR), systolic blood pressure (SBP), and body temperature. We used the closest set to the start date & time of antibiotic therapy (considering observations taken any time within 6 hours before or after the start of antibiotics). If two values were equally close to the start date & time of antibiotic therapy, we used the ‘worst’ value (highest RR, HR and body temperature, and lowest SBP).

*Limb temperature measurements:*

Measurements were taken using a thermal imaging camera that attaches to a smartphone (FLIR ONE® Gen 3 - Android USB-C (Teledyne FLIR, USA)).^8^ Measurements were taken at the point of maximal temperature on the affected limb and at the corresponding point on the non-affected limb, which also allowed for calculation of the limb temperature difference (affected minus unaffected limb temperature). One repeat measurement was taken from both the affected and unaffected limbs. Temperature readings were made approximately 10 minutes after removing any clothes or dressings. The device was held at room temperature for at least 10 minutes before readings were taken. Outlying repeated limb temperature measurements were removed based on the frequency distributions of their standard deviations.

*Patient-reported symptoms:*

Patients reported the severity of their pain (on movement and at rest) and swelling due to cellulitis daily on a 10-point verbal and numerical scale.

### Supplementary Tables

| **Sub-component** | **Score** | **Descriptors** | | |
| --- | --- | --- | --- | --- |
| **Acute colour change** | 0 | No or small* area, mild colour change |  |  |
|  | 1 | Small area, moderate colour change | Medium^†^ area, mild colour change |  |
|  | 2 | Small area, severe colour change (e.g. fiery and/or purple) | Medium area, moderate colour change | Large^‡^ area, mild colour change |
|  | 3 |  | Medium area, severe colour change (e.g. fiery and/or purple) | Large area, moderate colour change |
| **Warmth** | 0 | Small area, slightly warmer than contra-lateral side |  | No difference between sides |
|  | 1 | Small area, obviously warmer than contra-lateral side | Medium area, slightly warmer |  |
|  | 2 | Small area, hot compared to contra-lateral side | Medium area, obviously warmer | Large area, slightly warmer |
|  | 3 |  | Medium area, hot | Large area, obviously warmer |
| **Tenderness** | 0 | No tenderness | | |
|  | 1 | Tender on mild pressure | | |
|  | 2 | Tender on light pressure | | |
|  | 3 | Tender at rest | | |
| **Oedema** | 0 | No or small area, some oedema |  |  |
|  | 1 | Small area, obvious oedema | Medium area, some oedema |  |
|  | 2 | Small area, considerable oedema | Medium area, obvious oedema | Large area, some oedema |
|  | 3 |  | Medium area, considerable oedema | Large area, obvious oedema |
| **Ulceration** | 0 | Intact skin, no ulceration |  |  |
|  | 1 | Small ulcers, interdigital cleft | Total surface area <1x1 cm | Not deep |
|  | 2 | Ulcers/wounds | Total surface area >1x1 cm but <5x5 cm | Quite deep |
|  | 3 | Ulcers/wounds | Total surface area >5x5 cm | Very deep (up to bone) |
| **Discharge** | 0 | Dry: dry dressing, or no wound | Scab, thus dry |  |
|  | 1 | Moist: wound(s) barely leak | One dressing for two or more days is sufficient |  |
|  | 2 | Wet: dressings need daily changing | Dressings changed 2x/day, but dressings are not sated | Large surface area, or many areas, which are “Moist” |
|  | 3 | Leaking: 2 or more sated dressings per day | Moisture absorbing pad considerably wet | Large surface area, or many areas, which are “Wet” |
| **Blistering** | 0 | No blistering | Normal skin consistency |  |
|  | 1 | Recently drained blister |  |  |
|  | 2 | Small blister (max 2x2 cm) |  |  |
|  | 3 | Blister (larger than 2x2 cm) |  |  |

#### Supplementary Table 1. Cellulitis Severity Score first proposed by Hepburn *et al*.^7^ modified from Cranendonk *et al*.^1^

Note: 7 subcomponents (colour change, oedema, warmth, pain, ulceration, discharge and blistering) being scored on a 4-point scale (none = 0, mild = 1, moderate = 2, severe = 3), creating a total score between 0 and 21.

* Small = an area up to half of the lower leg or equivalent

† Medium = almost the entire lower leg, or part of the lower leg and one joint

‡ Large = the entire lower leg and entire foot, or the entire lower leg and two joints

Score not validated.

#### Supplementary Table 2. Univariable associations between acute clinical response variables and ‘hospital-attended recurrence’ or ‘any recurrence’ used to determine which variables to impute (threshold P<0.15)

| **Variables with P≥0.15 on both outcomes** | **Variables with P<0.15 on either outcome** | **Variables imputed*** |
| --- | --- | --- |
| **Physical examination†** | | |
| Days 0/1, 0, 1, 2, 3, 2/3, and change in length, width, and ‘area’ | . | . |
| Days 0/1, 0, 1, 2, 2/3, and change in colour | Day 3 colour | Days 0, 1, 2, and 3 colour |
| Days 0/1, 0, 1, 2/3, and change in warmth | Days 2 and 3 warmth | Days 0, 1, 2, and 3 warmth |
| Days 0/1, 0, 1, 2, 3, 2/3, and change in tenderness | . | . |
| Days 0/1, 0, 1, and 2 oedema | Day 3 and 2/3 oedema, and change in oedema | Day 2/3 oedema and change in oedema |
| Days 0/1, 0, 1, 2, 3, and 2/3 ulceration | Change in ulceration | Day 0/1 ulceration and change in ulceration |
| Change in discharge | Day 0/1, 0, 1, 2, 3, and day 2/3 discharge | Day 0/1 and 2/3 discharge |
| Days 0, 1, 2, 3, and change in blistering | Day 0/1 and day 2/3 blistering | Day 0/1 and day 2/3 blistering |
| Days 0/1, 0, and change in total Cellulitis Severity Score | Days 1, 2, 3, and 2/3 total Cellulitis Severity Score | Days 0, 1, 2, and 3 total Cellulitis Severity Score |
| **Limb temperature** | | |
| Days 0/1, 2/3, and change in affected limb temperature | Day 0, 1, 2, and 3 affected limb temperature | Day 0, 1, 2, and 3 affected limb temperature |
| Days 0/1, 1, 2 and 2/3 limb temperature difference | Day 3 and change in limb temperature difference | Day 3 and change in limb temperature difference |
| **Vital signs** | | |
| Days 0/1, 0, 1, 2, 3, 2/3, and change in respiratory rate | . | . |
| Days 0/1, 0, 1, 2, 3, and 2/3 heart rate | Change in heart rate | Day 0/1 heart rate and change in heart rate |
| Days 1, 2, and change in systolic blood pressure | Days 0/1, 0, 3, 2/3 systolic blood pressure | Day 0/1 and 2/3 systolic blood pressure |
| Day 2, and change in body temperature | Day 0/1, 0, 1, 3, and 2/3 body temperature | Day 0/1 and 2/3 body temperature |
| **Blood results** | | |
| Days 0/1, 0, 1, 2, 3, and 2/3 C-Reactive Protein, and change in C-Reactive Protein | . | . |
| Days 0/1, 1, 2, 3, and 2/3 neutrophil count | Change in neutrophil count | Day 0/1 and change in neutrophil count |
| Days 0, 1, and change in platelet count | Day 0/1, 2, 3, and 2/3 platelet count | Day 0/1 and 2/3 platelet count |
| Days 0/1, 0, 1, 2, 3, and 2/3 urea | Change in urea | Day 0/1 and change in urea |
| Days 0, 3, and change in albumin | Day 0/1, 1, 2, and 2/3 albumin | Day 0/1 and 2/3 albumin |
| **Patient-reported symptoms** | | |
| Days 0-7, 0/1, 2/3 patient-reported pain at rest, and changes between day 0 to day 1-7 | . | . |
| Days 0-7, 0/1, 2/3 patient-reported pain on movement, and changes between day 0 to day 1-7 | . | . |
| Days 0-7, 0/1, 2/3 patient-reported swelling, and changes between day 0 to day 2, 3, 4, 5, 6, and 7 | Change in patient-reported swelling between days 0 to 1 | Day 0 and change in patient-reported swelling between day 0 to 1 |

*We imputed variables with univariable P<0.15, including in imputations either absolute values from days 0-3, or absolute values from days 0/1 & 2/3, or change from baseline and the absolute baseline value, according to the specific variable with p<0.15.

† Variables represent Cellulitis Severity Score subcomponents, except length, width and ‘area’. ‘Area’ calculated by length multiplied by width/ ‘Change’ indicates day 0/1 to 2/3 difference unless otherwise indicated.

#### Supplementary Table 3. Univariable associations between acute clinical response variables and either ‘hospital-attended’ or ‘any recurrence’ with p<0.05

| **Variable** | **OR** | **95%CI** | **P** |
| --- | --- | --- | --- |
| Hospital-attended recurrence | | | |
| Day 0/1 blistering | 1.72 | 1.05-2.82 | 0.03 |
| Day 2/3 blistering | 2.25 | 1.42-3.57 | 0.001 |
| Day 2/3 discharge | 1.94 | 1.21-3.10 | 0.006 |
| Change in ulceration | 9.56 | 1.29-70.96 | 0.03 |
| Day 2 total Cellulitis Severity Score | 1.17 | 1.02-1.35 | 0.03 |
| Day 3 total Cellulitis Severity Score | 1.21 | 1.05-1.40 | 0.007 |
| Any recurrence | | | |
| Day 2/3 systolic blood pressure | 1.02 | 1.01-1.04 | 0.009 |
| Day 2/3 body temperature | 0.29 | 0.09-0.94 | 0.04 |

#### Note: All ORs are per unit higher. The full list of variables tested are those in the third column of Supplementary Table 2.

#### Supplementary Table 4. TRIPOD checklist: Prediction model development and validation

| Section/Topic | Item |  | Checklist Item | Page |
| --- | --- | --- | --- | --- |
| Title and abstract | | | | |
| Title | 1 | D;V | Identify the study as developing and/or validating a multivariable prediction model, the target population, and the outcome to be predicted. | 1 |
| Abstract | 2 | D;V | Provide a summary of objectives, study design, setting, participants, sample size, predictors, outcome, statistical analysis, results, and conclusions. | 2 |
| Introduction | | | | |
| Background and objectives | 3a | D;V | Explain the medical context (including whether diagnostic or prognostic) and rationale for developing or validating the multivariable prediction model, including references to existing models. | 3 |
|  | 3b | D;V | Specify the objectives, including whether the study describes the development or validation of the model or both. | 3 |
| Methods | | | | |
| Source of data | 4a | D;V | Describe the study design or source of data (e.g., randomized trial, cohort, or registry data), separately for the development and validation data sets, if applicable. | 3 |
|  | 4b | D;V | Specify the key study dates, including start of accrual; end of accrual; and, if applicable, end of follow-up. | 3&4 |
| Participants | 5a | D;V | Specify key elements of the study setting (e.g., primary care, secondary care, general population) including number and location of centres. | 3 |
|  | 5b | D;V | Describe eligibility criteria for participants. | 3 |
|  | 5c | D;V | Give details of treatments received, if relevant. | 4 |
| Outcome | 6a | D;V | Clearly define the outcome that is predicted by the prediction model, including how and when assessed. | 3 |
|  | 6b | D;V | Report any actions to blind assessment of the outcome to be predicted. | NA |
| Predictors | 7a | D;V | Clearly define all predictors used in developing or validating the multivariable prediction model, including how and when they were measured. | 3 |
|  | 7b | D;V | Report any actions to blind assessment of predictors for the outcome and other predictors. | NA |
| Sample size | 8 | D;V | Explain how the study size was arrived at. | 2 |
| Missing data | 9 | D;V | Describe how missing data were handled (e.g., complete-case analysis, single imputation, multiple imputation) with details of any imputation method. | 4 |
| Statistical analysis methods | 10a | D | Describe how predictors were handled in the analyses. | NA |
|  | 10b | D | Specify type of model, all model-building procedures (including any predictor selection), and method for internal validation. | NA |
|  | 10c | V | For validation, describe how the predictions were calculated. | 4&5 |
|  | 10d | D;V | Specify all measures used to assess model performance and, if relevant, to compare multiple models. | 4&5 |
|  | 10e | V | Describe any model updating (e.g., recalibration) arising from the validation, if done. | NA |
| Risk groups | 11 | D;V | Provide details on how risk groups were created, if done. | 4 |
| Development vs. validation | 12 | V | For validation, identify any differences from the development data in setting, eligibility criteria, outcome, and predictors. | 4 |
| Results | | | | |
| Participants | 13a | D;V | Describe the flow of participants through the study, including the number of participants with and without the outcome and, if applicable, a summary of the follow-up time. A diagram may be helpful. | supp |
|  | 13b | D;V | Describe the characteristics of the participants (basic demographics, clinical features, available predictors), including the number of participants with missing data for predictors and outcome. | 4 |
|  | 13c | V | For validation, show a comparison with the development data of the distribution of important variables (demographics, predictors and outcome). | supp |
| Model development | 14a | D | Specify the number of participants and outcome events in each analysis. | 4&5 |
|  | 14b | D | If done, report the unadjusted association between each candidate predictor and outcome. | NA |
| Model specification | 15a | D | Present the full prediction model to allow predictions for individuals (i.e., all regression coefficients, and model intercept or baseline survival at a given time point). | NA |
|  | 15b | D | Explain how to the use the prediction model. | NA |
| Model performance | 16 | D;V | Report performance measures (with CIs) for the prediction model. | 5 |
| Model-updating | 17 | V | If done, report the results from any model updating (i.e., model specification, model performance). | 5 |
| Discussion | | | | |
| Limitations | 18 | D;V | Discuss any limitations of the study (such as nonrepresentative sample, few events per predictor, missing data). | 6 |
| Interpretation | 19a | V | For validation, discuss the results with reference to performance in the development data, and any other validation data. | 5&6 |
|  | 19b | D;V | Give an overall interpretation of the results, considering objectives, limitations, results from similar studies, and other relevant evidence. | 5-7 |
| Implications | 20 | D;V | Discuss the potential clinical use of the model and implications for future research. | 7 |
| Other information | | | | |
| Supplementary information | 21 | D;V | Provide information about the availability of supplementary resources, such as study protocol, Web calculator, and data sets. | 7 |
| Funding | 22 | D;V | Give the source of funding and the role of the funders for the present study. | 7 |

#### Supplementary Table 5: Baseline characteristics, antibiotic treatment and microbiological results

|  | **External validation cohort (N=202)** | **Missing (%)** | **Development sample**  **(N=4,938)** | **SMD* validation vs development** |
| --- | --- | --- | --- | --- |
| Age (IQR) | 66 (51, 79) | 0 | 67 (51, 81) | 0.01 |
| Female (%) | 84 (42) | 0 | 2,297 (47) | 0.10 |
| White ethnicity (%) | 191 (95) | 8 (4) | 4,093 (83) | 0.13 |
| Other ethnicity (%) | 3 (1) |  | 150 (3) |  |
| **Cellulitis episode details** |  |  |  |  |
| Duration of symptoms prior to presentation (IQR) | 3 (1, 6) | 3 | ·· | ·· |
| Pre-existing lesion (%) | ·· | ·· | ·· | ·· |
| None | 85 (42) | 0 | ·· | ·· |
| Non-surgical trauma | 42 (21) | 0 | ·· | ·· |
| Ulcer | 32 (16) | 0 | ·· | ·· |
| Insect bite | 8 (4) | 0 | ·· | ·· |
| Animal bite | 4 (2) | 0 | ·· | ·· |
| Surgical site | 2 (1) | 0 | ·· | ·· |
| Other | 29 (14) | 0 | ·· | ·· |
| **Comorbidities** |  |  |  |  |
| Patient-reported previous cellulitis (%) | 105 (52) | 1 (0) | ·· | ·· |
| 1 prior episode | 46 (23) | ·· | ·· | ·· |
| 2 prior episodes | 26 (13) | ·· | ·· | ·· |
| $\geq$3 prior episodes | 33 (16) | ·· | ·· | ·· |
| Hospital-attended previous cellulitis (%) | 55 (27) | ·· | 229 (5) | **0.65*** |
| Venous insufficiency inc. venous ulcers (%) | 23 (11) | 0 | 210 (4) | 0.27* |
| Other ulcer disease**^†^** (%) | 43 (21) | 0 | 1,091 (22) | 0.02 |
| Lymphoedema (%) | 14 (7) | 0 | 147 (3) | 0.16 |
| Peripheral vascular disease (%) | 11 (6) | 0 | 427 (9) | 0.13 |
| Peripheral neuropathy (%) | 8 (4) | 0 | 209 (4) | 0.01 |
| Diabetes mellitus (%) | 42 (21) | 0 | 1,262 (26) | 0.11 |
| Congestive heart failure (%) | 25 (12) | 0 | 630 (13) | 0.01 |
| Liver disease (%) | 12 (6) | 0 | 261 (5) | 0.05 |
| Renal disease (%) | 21 (10) | 0 | 553 (11) | 0.03 |
| Cancer (%) | 8 (4) | 0 | 456 (9) | 0.21* |
| Immunosuppression (%) | 10 (5) | 0 | 277 (6) | 0.03 |
| Rheumatological disease (%) | 4 (2) | 0 | 242 (5) | 0.16 |
| Chronic pulmonary disease (%) | 34 (17) | 0 | 978 (20) | 0.08 |
| Myocardial infarction (%) | 4 (2) | 0 | 308 (6) | 0.22* |
| Charlson Comorbidity Index (IQR) | 0 (0, 10) | 0 | 4 (0, 14) | 0.16 |
| Elixhauser score (IQR) | 0 (0, 7) | 0 | 3 (0, 9) | 0.07 |
| **Vital signs** |  |  |  |  |
| Respiratory rate (IQR) | 18 (17, 20) | 18 (9) | 18 (16, 19) | 0.29* |
| Heart rate (bpm) (IQR) | 92 (81, 102) | 8 (4) | 85 (74, 97) | 0.39* |
| Systolic blood pressure (mmHg) (IQR) | 130 (119, 147) | 17 (8) | 130 (116, 145) | 0.11 |
| Body temperature (^o^C) (IQR) | 36.8 (36.5, 37.5) | 16 (8) | 36·7 (36·2, 37·3) | 0.33* |
| **Blood results** |  |  |  |  |
| Haemoglobin (g/L) (IQR) | 134 (121, 147) | 8 (4) | 127 (112, 140) | 0.42* |
| Platelets (×10^9^/L) (IQR) | 226 (179, 282) | 9 (5) | 250 (197, 318) | 0.24* |
| Neutrophils (×10^9^/L) (IQR) | 9.9 (7.0, 14.8) | 8 (4) | 8·0 (5·5, 11·2) | 0.43* |
| Eosinophils (×10^9^/L) (IQR) | 0.1 (0.0, 0.1) | 8 (4) | 0·08 (0·02, 0·18) | 0.07 |
| C-reactive protein (mg/L) (IQR) | 193 (48, 204) | 9 (5) | 64 (21, 144) | 0.42* |
| Urea (mmol/L) (IQR) | 6.0 (4.7, 9.1) | 9 (5) | 6·0 (4·4, 8·7) | 0.02 |
| Albumin (g/L) (IQR) | 39 (37, 42) | 73 (36) | 32 (28, 36) | **1.55*** |
| ALT (IU/L) (IQR) | 20 (14, 29) | 71 (35) | 19 (14, 29) | 0.03 |
| ALP (IU/L) (IQR) | 85 (67, 116) | 67 (33) | 86 (70, 113) | 0.04 |
| Bilirubin (µmol/L) (IQR) | 11 (7, 17) | 71 (35) | 10 (7, 15) | 0.16* |
| **Antibiotic therapy** |  |  |  |  |
| $\geq$2 antibiotic agents | 159 (79) | 0 | 2,751 (56) | **0.51*** |
| Length of (antibiotic) therapy, days (IQR) | 13 (10, 18) | 0 | 8 (6,11) | 0.40* |
| Days of (antibiotic) therapy, days (IQR) | 15 (10, 22) | 0 | 9 (7,13) | 0.30* |
| Beta-lactam allergy | 34 (17) | 0 | ·· | ·· |
| Admitted on antibiotics | 61 (30) | 0 | ·· | ·· |
| Duration of prior antibiotics, days (IQR) | 3 (2, 4) | 0 | ·· | ·· |
| Only treated with oral antibiotics | 13 (6) | 0 | ·· | ·· |
| **Microbiological results** |  |  |  |  |
| Swab, pus, or fluid sample^‡^ | 71 (35) | ·· | ·· | ·· |
| **Positive** | 33 (46) | ·· |  |  |
| **Monomicrobial (N=23)** | 23 (70) | ·· | ·· | ·· |
| MSSA only | 11 (48) | ·· | ·· | ·· |
| MRSA only | 3 (13) | ·· | ·· | ·· |
| Streptococci only | 3 (13) | ·· | ·· | ·· |
| Gram-negative bacilli only | 4/ (17) | ·· | ·· | ·· |
| Other | 2 (9) | ·· | ·· | ·· |
| **Polymicrobial (N=10)** | 10 (27) | ·· | ·· | ·· |
| MSSA | 9 (90) | ·· | ·· | ·· |
| MRSA | 0 | ·· | ·· | ·· |
| Streptococci | 6 (60) | ·· | ·· | ·· |
| Gram-negative bacilli | 5 (50) | ·· | ·· | ·· |
| Anaerobes | 1 (10) | ·· | ·· | ·· |
| Blood culture | 89 (44) | ·· | ·· | ·· |
| **Positive** | 12 (13) | ·· | ·· | ·· |
| **Monomicrobial (N=10)** | 10 (83) | ·· | ·· | ·· |
| MSSA only | 1 (10) | ·· | ·· | ·· |
| MRSA only | 0 | ·· | ·· | ·· |
| Streptococci only | 8 (80) | ·· | ·· | ·· |
| Gram-negative bacilli only | 1 (10) | ·· | ·· | ·· |
| **Polymicrobial (N=2)** | 2 (17) | ·· | ·· | ·· |
| Streptococci | 2 (100) | ·· | ·· | ·· |
| Gram-negative bacilli | 2 (100) | ·· | ·· | ·· |
| **Outcomes** |  | ·· | ·· | ·· |
| Hospital-attended recurrence within 90 days | 17 (8) | 0 | 436 (9) | 0.01 |
| Any recurrence within 90 days | 46 (23) | ·· | ·· | ·· |
| Length of stay (IQR) | 8 (4, 13) | ·· | 5 (2, 10)^¶^ | 0.18 |

ALT (alanine transaminase); ALP (alkaline phosphatase); bpm (beats per minute); IQR (interquartile range).

* Standardised mean difference (SMD) >0.20 some evidence, >0.5 moderate evidence of covariate imbalance between groups.

† Ulcer disease includes decubitus/pressure ulcers and ulcers of the skin not elsewhere classified.

‡ Samples from lower limb skin/ulcer/wound/lesion sent for microscopy, culture, and sensitivity.

#### Supplementary Table 6. Model performance of the BRRISC score extension using day-2/3 blistering

| **Outcome** | **Definition of previous cellulitis** | **Variable** | | **Beta coefficient** | **OR** | **95%CI** | **P** | **AUC** | **95%CI** |
| --- | --- | --- | --- | --- | --- | --- | --- | --- | --- |
| Hospital-attended recurrence | Patient-reported | BRRISC score (per unit) | | 0.52 | 1.68 | 1.25-2.26 | 0.001 | 0.84 | 0.75-0.92 |
|  |  | Day 2/3 blistering subscore vs. 0 | 1 | 1.30 | 3.68 | 0.64-21.28 | 0.2 |  |  |
|  |  |  | 2 | 2.20 | 9.04 | 1.51-54.20 | 0.02 |  |  |
|  |  |  | 3 | 2.75 | 15.6 | 2.81-86.83 | 0.002 |  |  |
|  | Hospital-attended | BRRISC score (per unit) | | 0.50 | 1.65 | 1.24-2.20 | 0.001 | 0.82 | 0.72-0.93 |
|  |  | Day 2/3 blistering subscore vs. 0 | 1 | 1.27 | 3.56 | 0.62-20.47 | 0.2 |  |  |
|  |  |  | 2 | 2.11 | 8.24 | 1.41-48.31 | 0.02 |  |  |
|  |  |  | 3 | 2.72 | 15.18 | 2.87-80.29 | 0.001 |  |  |

#### Supplementary Table 7. Net Reclassification Index (NRI)

| **Definition of previous cellulitis** | **Original BRRISC score risk categories** | | **Extended score**  **reclassified risk** | | | | | **% (N) of imputations**  **reclassified with** | | | **Net correctly reclassified (%)** |
| --- | --- | --- | --- | --- | --- | --- | --- | --- | --- | --- | --- |
|  |  |  | <5% | | 5 to 15% | >15% | | Increased risk | | Decreased risk |  |
| **Patient-reported** | **Patients with recurrence (N imputations=850)** | | | | | | | | | | **Event NRI** |
|  | <5% | | 0 | | 0 | 0 | | 34% (285) | | 0 | +34% |
|  | 5 to 15% | | 0 | | 115 | 285 | |  |  |  |  |
|  | >15% | | 0 | | 0 | 450 | |  |  |  |  |
|  | **Patients without recurrence (N imputations =9,250)** | | | | | | | | | | **Non-event NRI** |
|  | <5% | | 986 | | 59 | 152 | | 12% (1,103) | | 0 | -12% |
|  | 5 to 15% | | 0 | | 5,607 | 892 | |  |  |  |  |
|  | >15% | | 0 | | 0 | 1,554 | |  |  |  |  |
|  | **Summary NRI** | | | | | | | | | | **+22%** |
| **Hospital-attended** | **Patients with recurrence (N imputations =850)** | | | | | | | | | | **Event NRI** |
|  | <5% | 0 | | 0 | | 0 | 28% (235) | | 12% (98) | | +16% |
|  | 5 to 15% | 0 | | 165 | | 235 |  |  |  |  |  |
|  | >15% | 0 | | 98 | | 352 |  |  |  |  |  |
|  | **Patients without recurrence (N imputations =9,250)** | | | | | | | | | | **Non-event NRI** |
|  | <5% | 899 | | 143 | | 155 | 13% (1,185) | | 10% (937) | | -3% |
|  | 5 to 15% | 547 | | 5,065 | | 887 |  |  |  |  |  |
|  | >15% | 0 | | 390 | | 1,164 |  |  |  |  |  |
|  | **Summary NRI** | | | | | | | | | | **+13%** |

### Supplementary Figures

#### **Supplementary Figure 1. Flow diagram of participants**

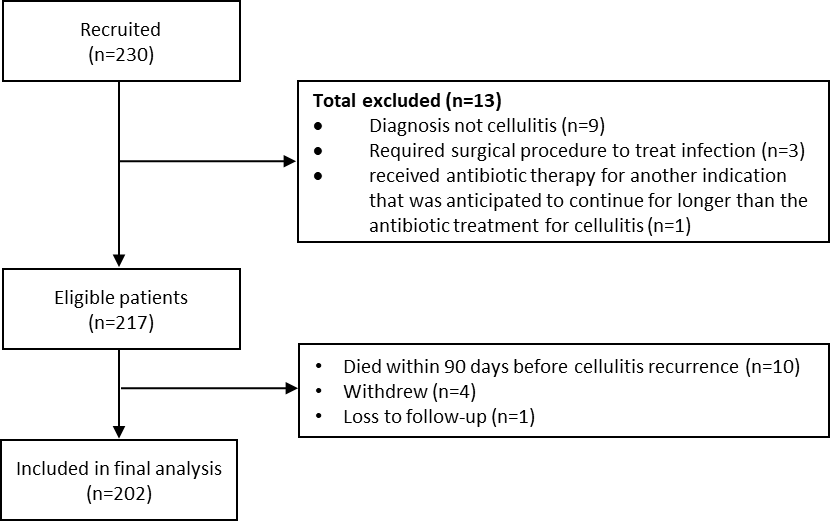

#### Supplementary Figure 2. Comparison of continuous original and imputed variables

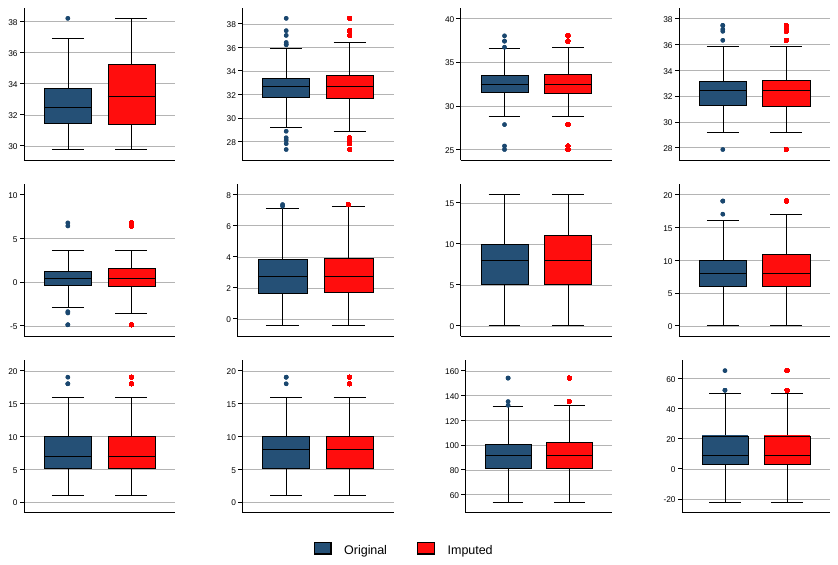

**Supplementary Figure 2. Comparison of continuous original and imputed variables**

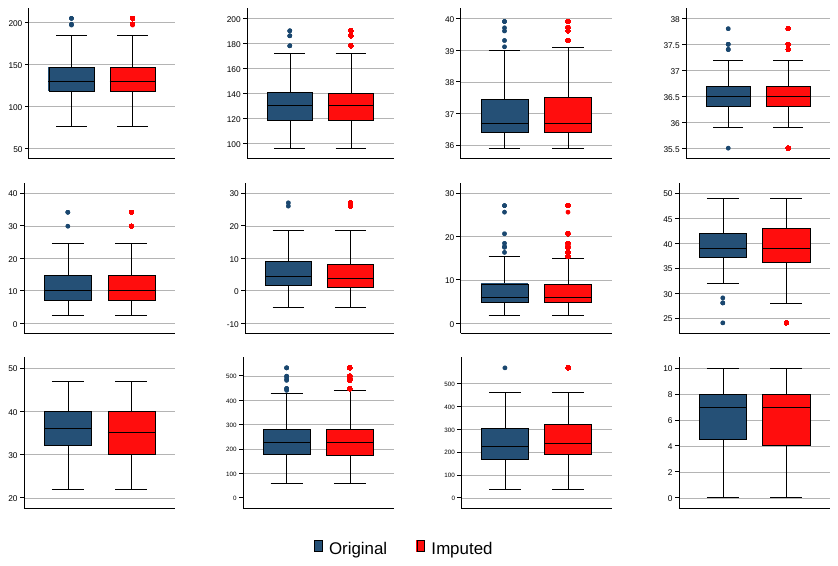

#### Supplementary Figure 3. Comparison of categorical original and imputed variables

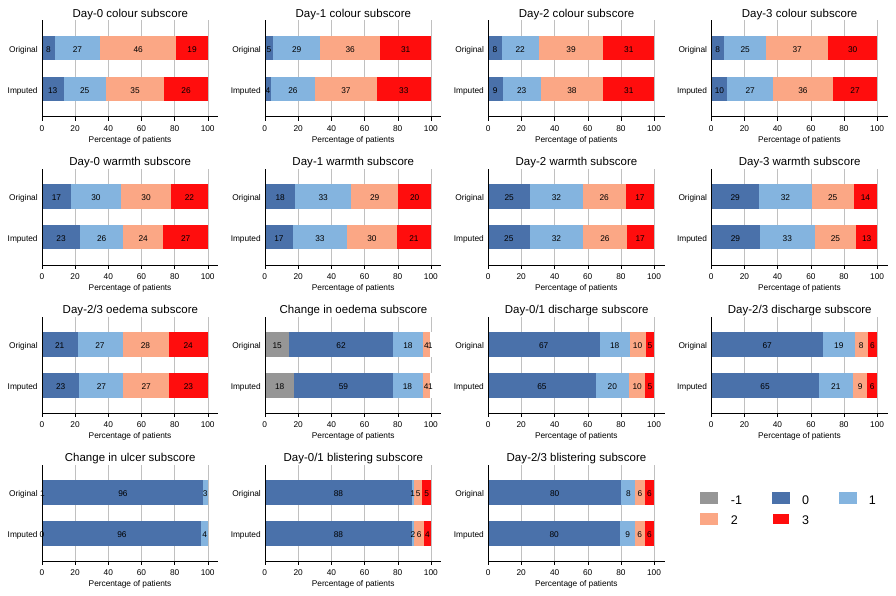

#### Supplementary Figure 4. Time to cellulitis recurrence

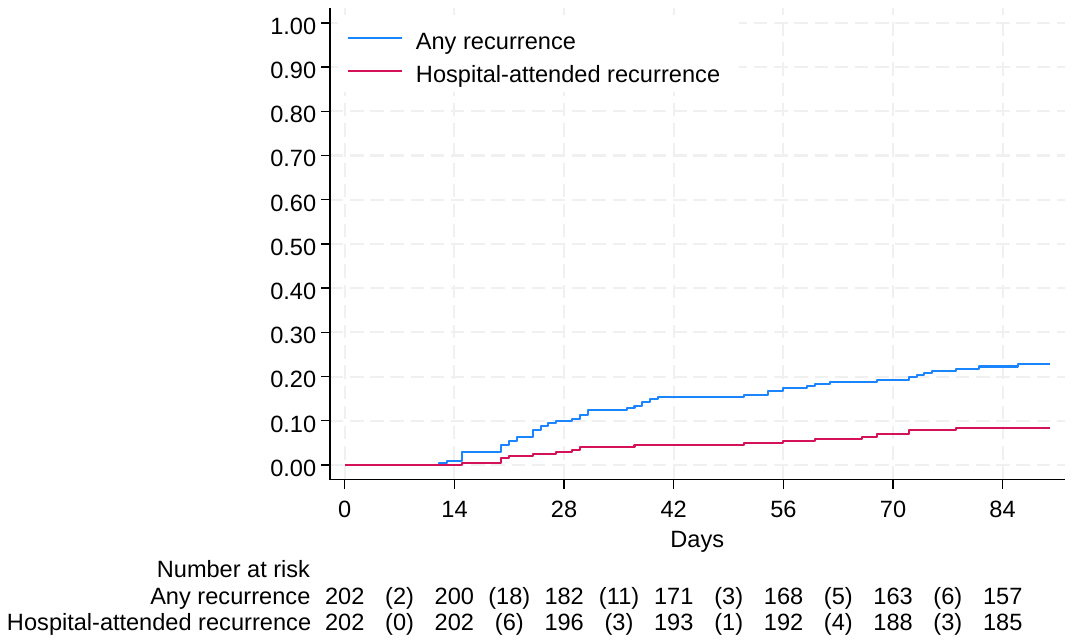

#### Supplementary Figure 5. Distribution of differences in C-indexes of BRRISC and extended score across the 50 imputed datasets

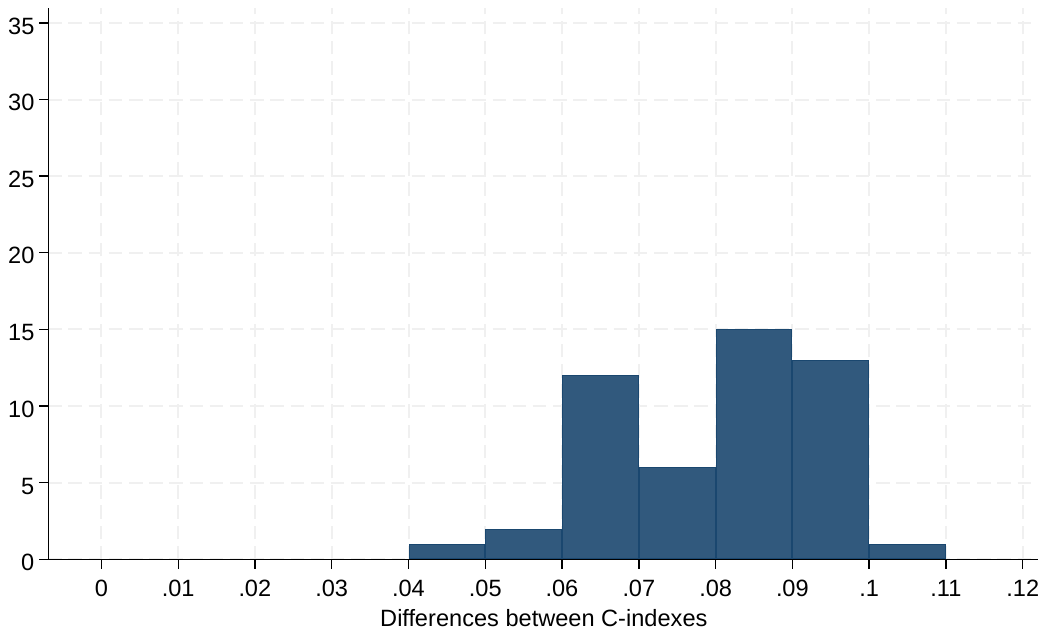

#### **Supplementary Figure 6. Distribution of P-values from tests for equality of C-indexes of BRRISC and extended score across the 50 imputed datasets**.

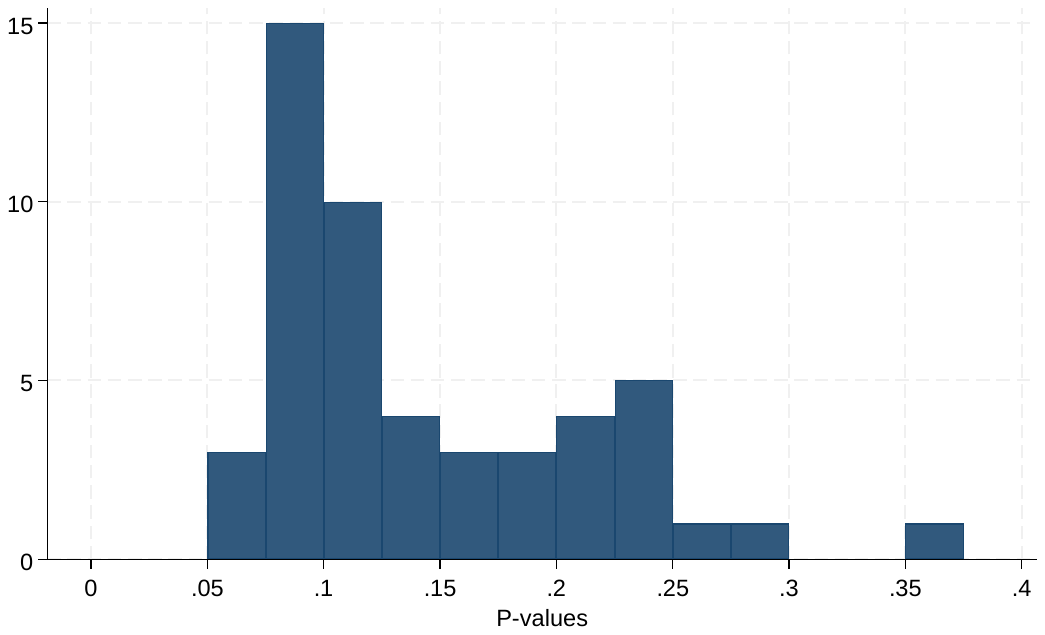
